# Is there hope for the Hajj? Using the SIR Model to Forecast COVID-19 Progression in the City of Makkah

**DOI:** 10.1101/2020.05.23.20105643

**Authors:** Alaa Mujallad, Haitham Khoj

## Abstract

**Background:** The Hajj is the largest annual gathering in the world, and it is a very important event for every Muslim. Makkah annually receives three million pilgrims who perform Hajj. Although precautionary measures have been taken in Saudi Arabia to slow down the spread of COVID-19, such as locking down the most affected cities, practicing social distancing, and applying the infection-control precautions, the number of cases has increased. The total confirmed number of cases in Makkah was 10,709 with 127 deaths as of May 16, 2020.

**Aims of the study:** Forecasting the COVID-19 progression in the city of Makkah will help the policymakers decide if the Hajj will be able to operate this year. Thus, to see a clear picture of the fight against COVID-19 for the economy and healthcare industry in Saudi Arabia, specifically in Makkah, the SIR model will predict COVID-19 progression in the city of Makkah.

**Method:** The Susceptible, Infected, and Recovered (SIR) model has been used to track the transmission dynamics and growth among the city of Makkah. The growth index was calculated, according to the data from March 16 until May 9. The estimated vital epidemiological parameters, such as forecasting works and transmission rates, were done.

**Result:** The data showed an interesting result about the peak of the disease progression. It is projected to occur around the 12th day after running the model. According to the model, the peak time will be around the 22^nd^ of May. Then, the number of cases will start to decrease.

**Conclusion:** Using the SIR model, the result predicts the disease progression peak and an estimated end of COVID-19 in the city of Makkah to help the policymakers decide if the Hajj will be able to operate this year.

---

Uncertainty about COVID-19 leaves people wondering about returning to their normal lives. Social distancing affects family gatherings and religious rituals. Mosques in Saudi Arabia have been closed to reduce the spread of COVID-19. Makkah is located in Saudi Arabia and hosts the largest annual Muslim ritual gatherings in the world, the Hajj. Muslims are waiting for this holy gathering with the hope that they can perform the Hajj, which will be around August 22^nd^, 2020. Forecasting the disease progression in the city of Makkah will help the policymakers to decide if the Hajj will be able to operate this year. Thus, to see a clear picture of the fight against COVID-19 for the economy and healthcare industry in Saudi Arabia, specifically in Makkah, the SIR model will predict COVID-19 progression in the city of Makkah.

Makkah is considered one of the holiest cities in Islam. Approximately eight in ten people around the world follow a religious group. Muslims are considered the second largest religious group worldwide after Christianity. There are 1.6 billion Muslims around the world 1. Makkah’s population is 2,042,000, without counting the large number of undocumented residents 2. Since the discovery of oil in Saudi Arabia, general development increased around 1960. Then, Saudi Arabia has become an attractive country in which to live. The Saudi way of life has adopted higher standards to boost the economy; the Saudi cities became developed, and new jobs were opened. Although Saudi Arabia has a strong economy, it is suffering from a shortage of a local workforce. Due to the economic difficulties in some Arab and South East Asian countries, Saudi Arabia was the best country to immigrate to work in for the last two decades 3.

Historically, pilgrims who came to Makkah brought with them many diseases such as cholera, plaque, and smallpox, which frequently occurred in over-crowded places. The holy places are frequently packed with people. That’s why many times epidemics happened. In the nineteenth century, Makkah became the station of cholera between Bangal and Egypt. Cholera arrived in Makkah from countries with cholera pandemics such as Java and Singapore, and it killed 15,000 people out of 90,000. The effects- of cholera traveled with pilgrims to their own countries, causing 60,000 deaths in Egypt 4. Due to the disease that travelled with the pilgrims, Saudi Arabia’s health officials issued visa requirements for entry for Hajj and Umrah. The requirements include vaccination against pandemic influenza A (H1N1), yellow fever, bubonic plaque, meningococcal meningitis, and poliomyelitis 5. Applying these requirements reduced the spread of diseases among the pilgrims. However, COVID-19 has no approved treatment and vaccination to prevent its spreading 6.

Containing COVID-19 is not an easy thing to do across Saudi Arabia, especially in the city of Makkah, due to the undocumented immigrants who have been living there for decades. The undocumented immigrants in Saudi Arabia are categorized into four groups. The first group came for Hajj, or Umrah, and extended their stay in Makkah without permission. The second group came for a working visa and stayed longer after violating their work contract. Third are the people who were smuggled from the neighboring countries such as Yemen. The fourth group is the children who were born from illegal immigrants; their status in Saudi Araba was considered illegal 7.

The government’s actions were effective in preventing the spread of the disease among those who violated their stay in Saudi Arabia. The Saudi government announced that all undocumented immigrants would be screened for COVID-19, and provided the treatment needed for them for free without asking about their legality of stay. Moreover, the health officials targeted the areas with high numbers of undocumented immigrants to test the largest amount of people there. Makkah now has the most COVID-19 patients among the Saudi cities, but with these actions from the government, the number is expected to go down 8.

COVID-19 is highly contagious, which means it has the ability to spread very fast. The total confirmed cases in Saudi Arabia is 52,016 with 302 deaths (WHO) as of May 16, 2020. Although precautionary measures have been taken in Saudi Arabia, such as locking down the most affected cities, practicing social distancing, and applying the infection control precautions (MOH), the number of cases has increased. The total confirmed cases in Makkah is 10,709 with 127 deaths (MOH).

Investigating various types of epidemic models, the main purpose is to understand the mechanism of disease progression. There are many models for forecasting, and the SIR model is one of the models that has a proven history to predict many infectious diseases. Susceptible, Infected, and Recovered (SIR) is a frequently used model that tracks the transmission dynamics and growth amongst the population. The usage of SIR model is to estimate vital epidemiological parameters, such as forecasting works and transmission rates 9. It is supportive to use a mathematical procedure that has dealt with various infectious diseases and apply it to COVID-19 to have an evidence basis for forecasting.

## Method

Based on the nature of COVID-19 and the characteristic of the virus, SARS-CoV-2 that causes the disease, a SIR model is a good fit to forecast the end period of COVID-19. The growth index will be calculated according to the data from March 16 until May 9. Susceptible, Infected, and Recovered are the main state variables of the model. The susceptible will be the whole population of the city of Makkah because this disease is highly contagious, closer to the 2000000. This data was taken from an authoritative source (Ministry of health). The infected and the recovered are secondary data that have been taken from (Ministry of health).

The SIR model first introduced by Kermack and Mckendrick (1927) 10, where they have three components susceptible, infected, and recovered as the disease spread. In our study we employ a SIR model to predict the peaks of cases before they decrease over time. The importance of our study is to give a prediction to all Muslim people all over the world whether they can complete one of their five pillars of Islam (Hajj). Additionally, according to the Ministry of Tourism, 75% of hotel rooms are in Makkah and Medina, which economically have a negative impact on tourism sectors and increase the unemployment rate. However, a study found that during a flu pandemic it is cost efficient to lockdown travel if death rate is high or above average 11. Today, the number of cases in Makkah is still increasing at a sustainable rate, because of the preventive measure that health officials are taking to control the spread of the virus. While everyone is hoping to an end to COVID-19 or at least control the number of cases, it’s important to forecast and predict the length of time to lessen the lockdown and open gradually.

So, a population of N in our study divided to (S) susceptible, (I) infected, and (R) recovered

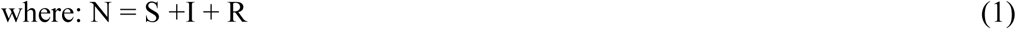

So, where rewrite it as a function of time as follows:

S = S(t) number of susceptible individuals over time,

I = I(t) number of infected individuals over time, and

R = R(t) number of recovered individuals over time.

Thus, Susceptible differential equation

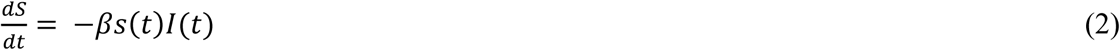

The Recovered differential Equation. R (t)

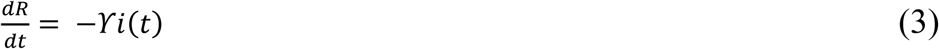

At the initial condition at initial time {S (0), I (0), R (0)} where parameter β = 1.7 per day is the infection transmission rate and parameter ϒ = .32 the recovery rate.

Parameter β represent the rate of Susceptible who get infected over time, and parameter ϒ represent the recovery rate whose been infected. By running the simple SIR model we can forecast when is the peak time of COVID-19 in Makkah over time where t = 70.

## Results

In this section, the results of the SIR calculations were presented. The time period was 70 days as the study in China calculated their sample of 70 days 12. We have S (0) = .9999 of the population is susceptible and starting infected at I (0) = 0.0001, which was a percent of population infected, and recovery rate is zero R (0) = 0. We compared our findings with China’s cases, and it presented similar results to the constant parameters for the SIR model that has been used to forecast the epidemic COVID-19 in China. The following table 1 shows the state variable parameters for the SIR model in the city of Makkah, Saudi Arabia. It is important to declare that Saudi Arabia is implementing a lockdown of all activities and ordering individuals to stay at home which we did not consider in our study.

**Table 1:** Summary of constant parameters for SIR model to forecast the epidemic COVID-19 in Makkah.

|  |  |
| --- | --- |
| Place | Makkah |
| Population | 2000000 |
| Susceptible rate | .9999 |
| Infection rate | .0001 |
| Recovered | 0 |

The data presented in graph 1 is the daily number of cases reported in Saudi Arabia, and the daily number reported in Makkah from 3/16/2020 to 5/09/2020. Both curves indicate the increasing number over time, with 673 being the daily average of infected people in Saudi Arabia and the average daily cases in Makkah is 145.

**Graph 1:**
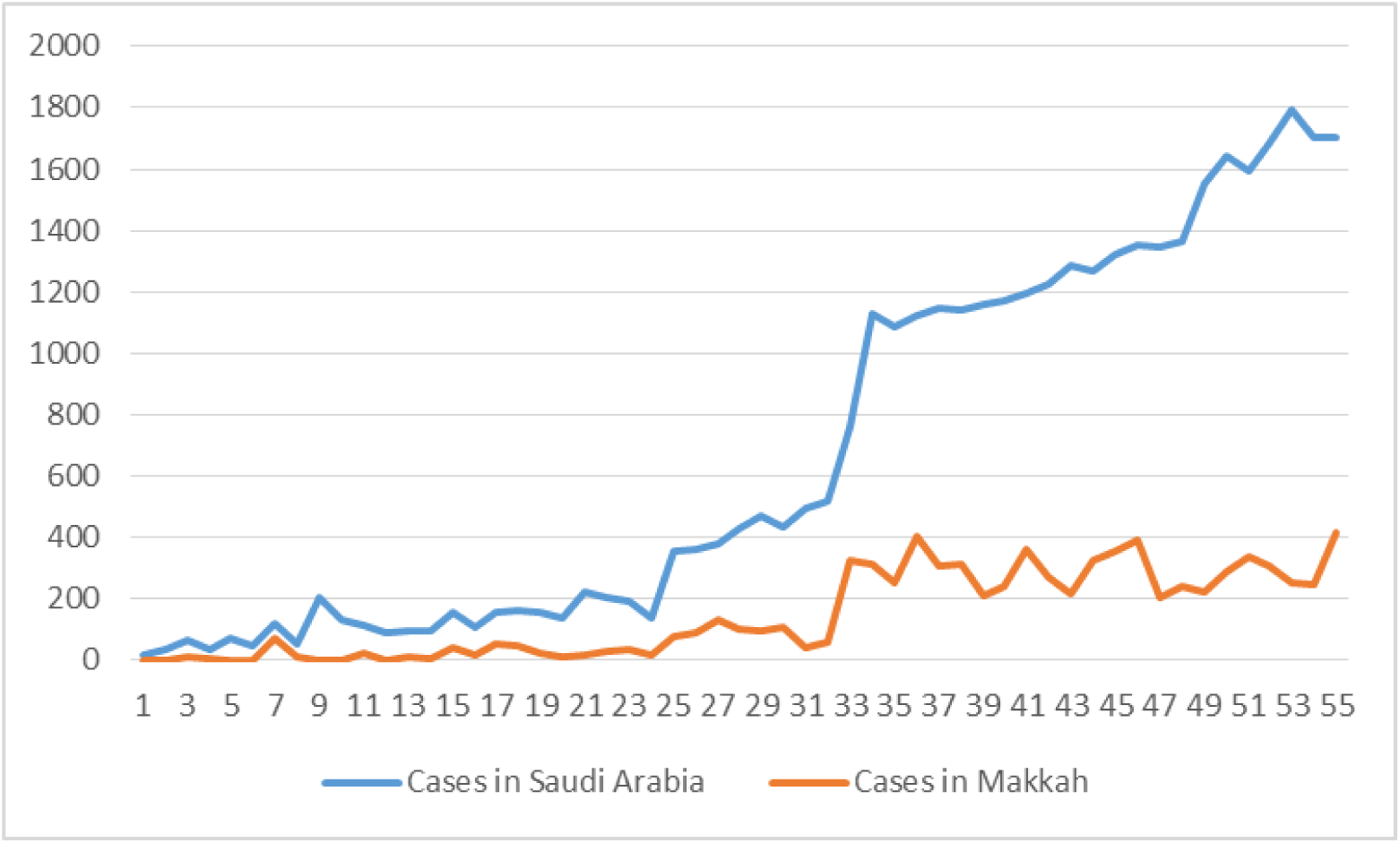
Number of daily cases in Saudi Arabia VS Makkah

**Graph 2:**
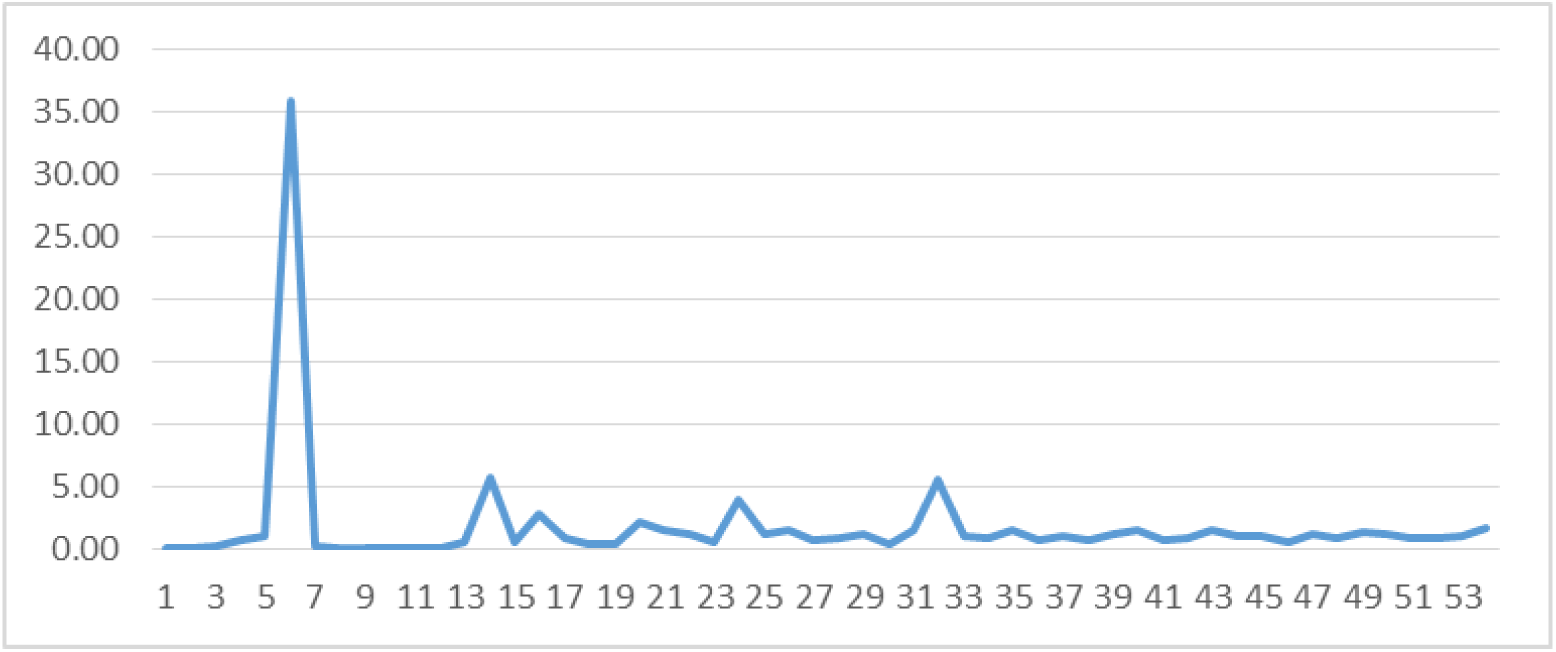
Growth index of Daily Cases

To have a better understanding of the speed of COVID-19, we calculate the growth index. Graph 3 shows our calculation of the growth index of COVID-19 from the first reported date of 3/16/2020 to 5/9/2020, and it is clear that we had a big spike after the 5th day of the first reported case. On average, the growth index is 1.7.

Figure 1 shows the dynamics of the three variables S, I, and R. We observe that the curve of susceptible is decreasing over time due to the assumption that the birth rate = 0. Also, we assumed that once susceptible people are infected, they will never return to a state variable of susceptible. The infected curve shows that the number of infected people will reach the peak in 15 days before it starts to decrease. The recovery curve starts low before it increases dramatically as the number of recovered individuals accumulates and rises above the infected curve. The intersect between the recovery curve and infected curve assumingly will be in 18 days, which is the beginning of June. The data showed an interesting result about the peak of the disease progression. It is estimated to occur around the 15th day after running the model. According to the model, the peak time will be around the 22^ed^ of May. Then, the cases will decrease until the 30^th^ day, which is around the 15^th^ of June.

**Figure 1:**
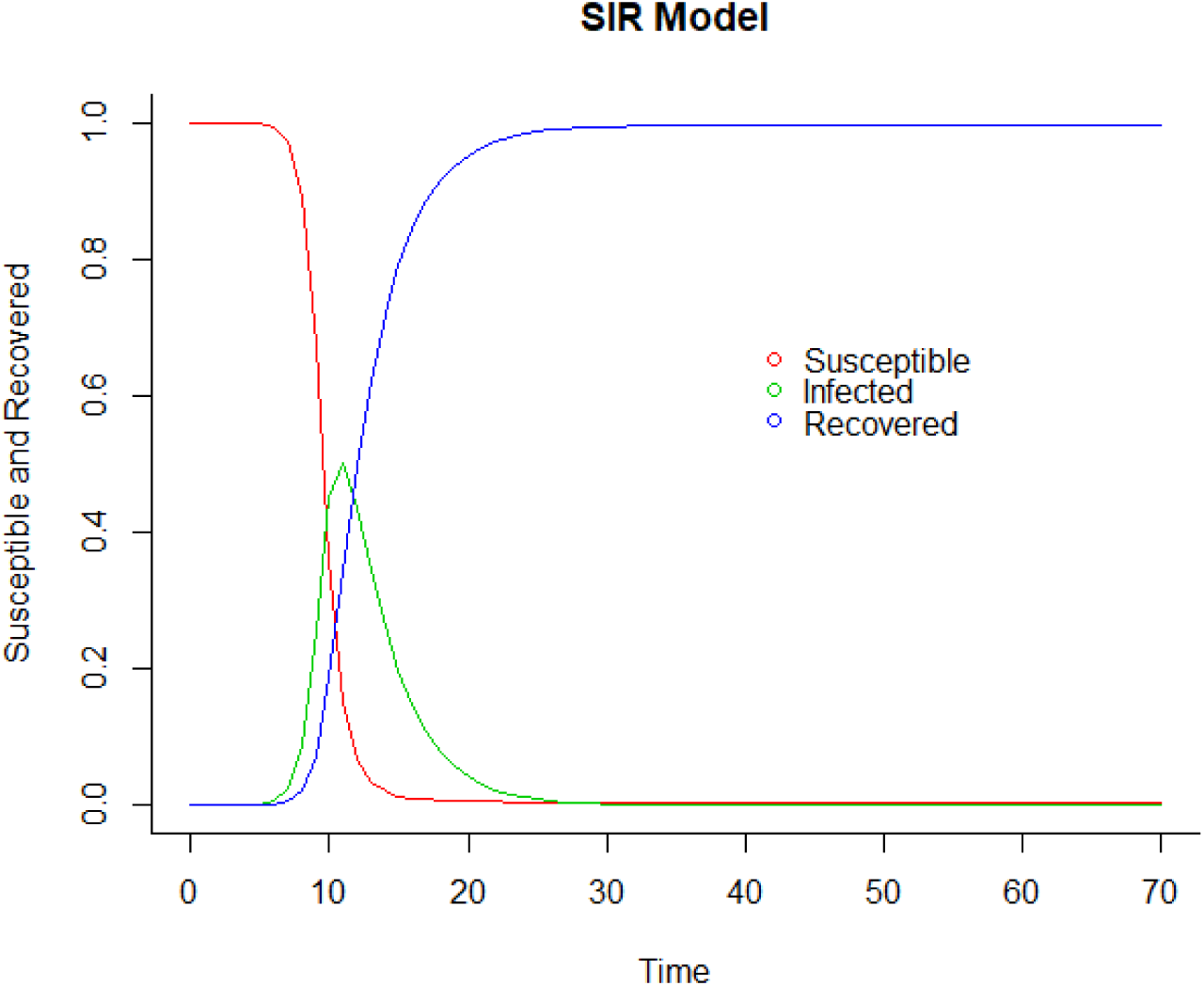
The SIR Model to Forecast COVID-19 Progression in the City of Makkah

## Discussion

This study has been done in a very important time in the ongoing COVID-19 pandemic. The main goal of the study was estimating the disease progression in the city of Makkah and to help the policy makers to decide if the Hajj will be held this year. One of the important features of a SIR model is the feature of identifying simultaneously with model parameters, which can detect and forecast positive infected people of the ϒ factor that relates to the actual number of infected individuals in the sample population 13. For the accumulated data of Saudi Arabia, the recovery rate is 32%, and the real mortality rate of the infection is .5% as of May 9 (Ministry of Health). The data covers 70 days since the emergence of the first infected person in a city of Makkah, Saudi Arabia, which was on March 16th (Ministry of Health). The parameters have been used according to this data. The model and the parameters allowed us to forecast the time needed to predict the end time of COVID-19 in Makkah or at least when the curve would be flattened. The other countries that had a decline took around three months. So, the time was estimated around 70 days. An important note about the study that it is did not consider Saudi Arabia is imposing prevention measures on the time period we are covering. Another important note is that on the 22^nd^ of March the number jumped significantly on the growth index, which can be explained by the 72 reported cases at a hotel in Makkah (Ministry of Health). The findings provide significant outcomes for health authorities to ensure the availability of all resources that they might need. Also, it helps them to forecast when the curve will flatten and apply their economic and health polices accordingly. Figure 1 predicted that the curve will flatten in 20 days within the constraint that nothing changes. As of the 27th of February, Saudi Arabia stopped Umrah visas, which was an early prevention to slow down the spread of the virus and decrease the number of infected people.

## Conclusion

The economy and the healthcare industry have been most affected by this pandemic. Solidarity is now needed in the fight against COVID-19 to resume to the normal life, attend family gatherings, and perform religious rituals. Using the SIR model, the result predicts the disease progression peak and an estimated end of COVID-19 in the city of Makkah to help the policymakers decide if the Hajj will be able to operate this year.

## Data Availability

ministry of health in Saudi Arabia

## Notes

### Competing Interest Statement

The authors have declared no competing interest.

### Funding Statement

no funding

### Author Declarations

It is a secondary data

## References

1. Bock, D. L., & Del Rosario, M. (2014). The table briefing: Ministering to hospital patients. Bibliotheca Sacra, 171(682), 226–232.

2. World Population 2020

3. Algahtany, M., L. Kumar, and H. Khormi. 2016. Are immigrants more likely to be involved in criminal activity in Saudi Arabia? Open Journal of Social Sciences 403: 170–186. https://doi.org/10.4236/jss201643023.

4. Henderson RJ. Problems of pilgrimages. Postgrad Med J 1975; 51:845–847. [PMC free article] [PubMed] [Google Scholar]

5. WHO. Health conditions for travellers to Saudi Arabia for the pilgrimage to Mecca (Hajj). Wkly Epidemiol Rec 2009; 84:477–480. [PubMed] [Google Scholar]

6. Ning L, Liu L, Li W, et al. Coronavirus (SARS-CoV-2) infection in a renal transplant recipient: case report. Am J Transplant. 2020

7. Alsharif, Fahad L. City of Dreams, Disappointment and Optimism: The Case of Nine Communities of Undocumented African Migrants in the City of Jeddah.

8. Ministry of Health of Saudi Arabia (MOH)

9. W.-K. Ming, J. Huang, C. J. P. Zhang, Breaking down of the healthcare system: Mathematical modelling for controlling the novel coronavirus (2019- nCoV) outbreak in wuhan, china doi:10.1101/2020.01.27.922443. URL https://doi.org/10.1101%2F2020.01.27.922443

10. Kermack, W. O., and McKendrick, A. G. (1927). Contributions to the mathematical theory of epidemics, part i. Proceedings of the Royal Society of Edinburgh. Section A. Mathematics. 115 700–721.

11. J’er^ome Adda. Economic activity and the spread of viral diseases: Evidence from high frequency data. Quarterly Journal of Economics, 131(2):891–941, May 2016. doi:10.1093/qje/qjw005.

12. Pan A, Liu L, Wang C, et al. Association of public health interventions with the epidemiology of the COVID-19 outbreak in Wuhan, China. JAMA. Published online April 10, 2020. doi:10.1001/jama.2020.6130

13. G.C. Calafiore, C. Novara, C. Possieri A modified SIR model for the COVID-19 contagion in Italy (2020) arXiv preprint arXiv:2003.14391.

14. Ranney, Megan L., Valerie Griffeth, and Ashish K. Jha. 2020. “Critical Supply Shortages – The Need for Ventilators and Personal Protective Equipment during the Covid-19 Pandemic.” The New England Journal of Medicine, March 25. Available at: https://www.nejm.org/doi/full/10.1056/NEJMp2006141.

15. Zhan M, Qin Y, Xue X, Zhu S. Death from Covid-19 of 23 Health Care Workers in China. N Engl J Med. 2020 Apr 15. doi: 10.1056/NEJMc2005696.

16. Fernandes, Nuno, Economic Effects of Coronavirus Outbreak (COVID-19) on the World Economy (March 22, 2020). Available at SSRN: https://ssrn.com/abstract=3557504 or http://dx.doi.org/10.2139/ssrn.3557504.

